# Neural Mechanisms Underlying Attention Control In Relation To Anxiety And Depressive Symptoms

**DOI:** 10.1101/2023.11.10.23298383

**Authors:** Raye Fion Loh, Savannah Siew Kiah Hui, Junhong Yu

## Abstract

Poor attention control has been implicated in the development of anxiety and depression-related disorders and it is a key diagnostic criterion. This study aims to understand the possible neural mechanisms behind this.

191 German participants aged 20-80 were assessed on their level of attention control, depression and anxiety as part of the Leipzig Study for Mind-Body-Emotion Interactions. Network-based statistics were applied to their resting-state functional connectivity (rsFC) data to identify networks positively and negatively associated with attention control. Mediation analyses were then performed with these two networks as mediators.

Attention control correlated negatively with both anxiety and depression. The frontoparietal- or dorsal attention-somatomotor connections featured prominently in the attention control-positive network (ACPN). This network correlated positively with attention control, and negatively with both anxiety and depression. The attention control-negative network (ACNN) was largely represented by the ventral attention- or dorsal attention-visual connections. The ACPN was a significant and partial mediator between attention control and anxiety and a complete mediator for the relationship between attention control and depression. These findings could prove useful as neuroeducation in anxiety- and depression-related disorders, and as evidence for attention-based therapy.

## 1. Introduction

Anxiety disorders are common psychiatric disorders and major causes of disability and mortality throughout the world. Despite its adaptive benefits, anxiety becomes pathological when an individual starts to experience intense anxiety far beyond the expected levels for the situation, leading to excessive worry, hypervigilance, physiological arousal, and avoidance behaviours [1], depleting mental resources, reducing cognitive efficiency and negatively influencing daily functioning and quality of life [2, 3].

Depressive disorders are also highly prevalent and are characterised by emotional, cognitive, and physical symptoms [4]. Patients with depressive disorder have been frequently reported to present with cognitive impairment in several cognitive domains [5, 6] such as executive function [e.g. 9-12], episodic memory [e.g. 7, 13-17], semantic memory [e.g. 12, 18-22], visuo-spatial memory [e.g. 23, 24], and information processing speed [e.g. 6, 12, 17, 20-22]. Anxiety and depression symptoms often overlap significantly [25], which may point to possible underlying vulnerabilities that result in continued symptom perpetuation. One such underlying vulnerability may be poor attention control. Several depression models [26–28] have identified deficiencies in attention control and recurrent negative thinking (i.e., rumination) as important elements in the development and maintenance of negative affect. Therapies effective at reducing anxiety from baseline levels include those that support active goal-focused attention and flexible cognitive control, particularly inhibitory control [29]. This similarly suggests poor attention control is implicated in anxiety symptoms. The frequent co-occurrence of attention control deficits and anxiety symptoms could perhaps be explained by the possibility that both implicate the same resting-state networks. Resting-state functional connectivity (rsFC) can be quantified by the degree of temporal co-activation of spontaneous fMRI signals between various brain areas in the absence of a perceptual or behavioural task [30], and is helpful for measuring mind-wandering states such as attention control [31].

Hence, in the current study, we hypothesise that certain resting-state networks mediate the relationship between attention control and depression/anxiety in the non-clinical population. Few brain imaging studies have investigated this correlation in non-clinical populations. Even though these sub-clinical anxiety-related and depressive disorders are less understood as compared to their clinically significant counterparts [32], they are still prevalent [33, 34], often being associated with lower levels of quality of life [34–36], increased use of health services [34, 37], increased economic costs [33, 34] and higher mortality rates [34, 38]. More importantly, they are often early warning markers of subsequent clinical diagnosis [33, 38–40]. In addition, there have been few studies investigating the effect of attention control on the presentation of anxiety- and depression-related symptoms across the lifespan. Most studies mentioned above recruited participants who were young adults. However, anxiety and depression symptoms are not limited to young adults; rather, they are applicable even to the elderly [41].

## 2. Material & Methods

### 2.1 Participants

Datasets from an openly accessible anonymized database, the Leipzig Study for Mind-Body-Emotion Interactions project [42] and an additional follow-up project [43] were used in this study and details regarding recruitment and study procedure can be found there. This pThe study protocol was approved by the University of Leipzig’s medical faculty’s ethics committee (reference number 097/15-ff) [43]. The database was accessed on January 8, 2022, and there was no available access to identifying information.

Data from 13 subjects were eventually excluded (12 due to significant head motion during rs-fMRI scans, 1 due to missing data), leaving a total of 191 native German-speaking participants (92 females, 99 males). The specific age of each participant was not supplied in the dataset; instead, the age of each person is expressed in 5-year bins as shown in Figure 1.

**Figure 1.**
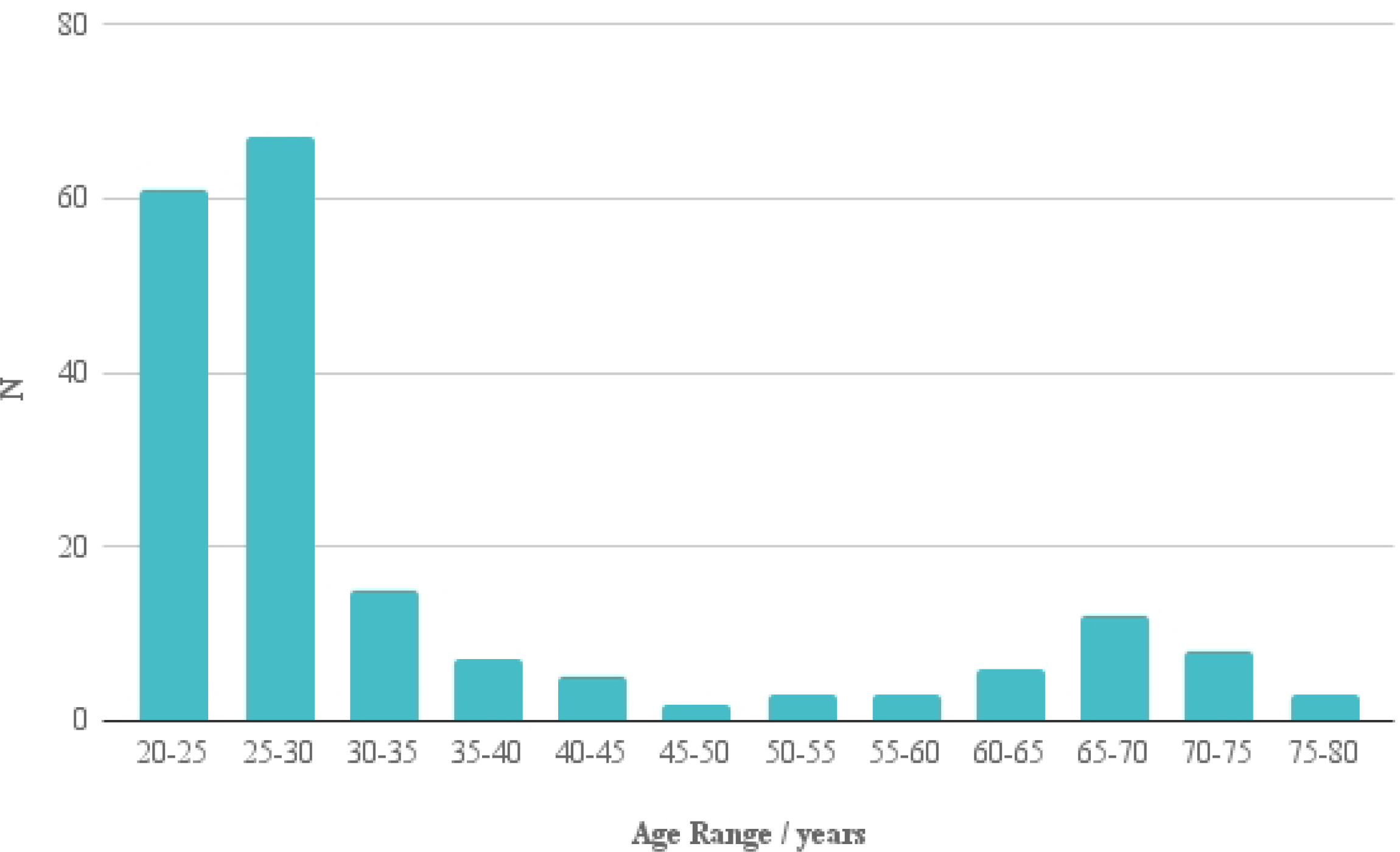
Summary of the Age Range of Par.

### 2.2. Measures

The Attention Control Scale [ACS] [42] was used in this study to evaluate individual differences in attention control. It is a self-report questionnaire with 20 items and it measures an individual’s ability (a) to focus, (b) to orientate attention between tasks and (c) to control thought flexibly. Participants responded on a 4-point Likert scale (1 = almost never; 2 = sometimes; 3 = often; 4 = always), where a higher score reflected greater attention control ability.

The Hospital Anxiety and Depression Scale [HADS] [45] was employed in this study to assess subclinical tendencies of depression and anxiety. It consists of 14 questions and two subscales measuring (a) anxiety [HADS.A] and (b) depression [HADS.D] across the past week. Each subscale has seven items and it is scored on a 4-point Likert scale (e.g., 1 = most of the time, 4 = never). A higher score represented higher anxiety and depression symptoms.

### 2.3. fMRI acquisition

The full fMRI acquisition details may be obtained from [42, 43]. Magnetic resonance imaging (MRI) was done using a 3 Tesla scanner equipped with a 32-channel head coil. Magnetization Prepared 2 Rapid Acquisition Gradient Echoes (MP2RAGE) procedure was used to collect T1-weighted images (TE=2920ms; TR=5000ms; TI1=700ms; TI2=2500ms; FOV=256mm; 176 sagittal slices; voxel size=1mm isotropic). T2-weighted gradient echo planar imaging (EPI) multiband BOLD technique (TR = 1,400 ms; TE = 30 ms; 64 axial slices; matrix = 88 x 88; voxel size = 2.3mm isotropic) was used to acquire the resting-state fMRI (rs-fMRI) volumes. Participants were told to stay awake and lie motionless with their eyes open while staring at a low-contrast fixation cross.

### 2.4. Data pre-processing

The T1 structural images were preprocessed with FreeSurfer 7.2.0 using the default recon-all options. Non-brain tissue was removed using a hybrid watershed or surface deformation procedure [46], automated Talairach transformation, segmentation of subcortical white matter and deep grey matter volumetric structures (including the hippocampus, amygdala, caudate, putamen, ventricles) [47, 48], intensity normalisation [49], tessellation [50, 51]. When the cortical models were finished, they were registered to a spherical atlas based on individual cortical folding patterns to match cortical geometry across participants [52], and the cerebral cortex was divided into units based on gyral and sulcal structure.

fMRIPrep 20.2.5 was used to preprocess the resting fMRI volumes [53]. Slice time was adjusted using 3dTshift from AFNI [54] and the motion was corrected using MCFLIRT [55]. Following this, co-registration to the matching T1w was performed using boundary-based registration [56] with 9 degrees of freedom, using bbregister from freesurfer. Motion correction transformations, BOLD-to-T1w transformations, and T1w-to-template (MNI) warps were combined and applied in a single step with antsApplyTransforms and Lanczos interpolation.

Following that, using the load confounds package (https://github.com/SIMEXP/load confounds), these preprocessed volumes were denoised by regressing out 6 motion parameters, the average signal of white matter and cerebrospinal fluid masks, global signal and their derivatives, as well as cosines covering the slow time drift frequency band. Scrubbing was used to reduce the effects of excessive head motion [57]. After that, the volumes are smoothed with a 5mm FWHM kernel and sent through a 0.1Hz low-pass filter. Finally, the brainnetome atlas [58] was used to divide the whole brain into 246 anatomical areas that corresponded to network nodes. Participants with significant head motion are omitted from the analysis if more than 20% of their rs-fMRI volumes are over the high motion limit (relative RMS > 0.25).

### 2.5. Statistical analysis

#### 2.5.1. Network-based statistics

First, network-based statistics were carried out on the rsFC matrices to obtain edges correlated with attention control. The selection thresholds were set at *p* < .01 and *p* < .05 at the edge and network levels respectively. Significant edges were separated into those positively correlated with attention control, labelled as the attention control-positive network (ACPN), and those negatively correlated with attention control, labelled as the attention control-negative network (ACNN). Connectivity scores for each participant were calculated for both positive and negative networks based on the connectivity strength of each network such that the greater the network score, the greater the connectivity strength of the networks. These analyses were performed in R (version 4.1.0) using the *NBR* package [59].

#### 2.5.2 Mediation analysis

Subsequently, the ACPN and ACNN scores were used as individual mediators of the connection between attention control and anxiety, as well as attention control and depression. This analysis was conducted in R with the mediation package [60], employing a statistical significance of *p* < 0.05, with quasi-Bayesian confidence intervals with 1000 Monte Carlo simulations.

#### 2.5.3. Data and code availability

The preprocessed rsFC matrices, behavioural scores of the participants studied, and the R code for the analysis may be found at *osf.io/f2qd6*.

## 3. Results

### 3.1. Network-based analysis

The attention control-related brain connection patterns are shown in Figure 2. The edges were classified into seven brain networks [61]. Attention control is positively associated with increased somatomotor-frontoparietal and somatomotor-dorsal attention connectivity. Strong positive connections can be observed between the frontoparietal and somatomotor networks, as well as between the dorsal attention and somatomotor networks. More specifically, there were stronger connections between the primary motor cortex and parts of the dorsal and medial prefrontal cortex. Attention control is negatively associated with decreased visual-ventral and visual-dorsal attention connectivity. Strong negative connections can be observed between dorsal attention and visual networks, as well as between ventral attention and visual networks, between the caudal dorsolateral region and the left inferior occipital gyrus, left occipital polar cortex, left medial superior occipital gyrus and left lateral superior occipital gyrus as well as opercular area 44 and right inferior occipital gyrus, right medial superior occipital gyrus, right rostral lingual gyrus.

**Figure 2.**
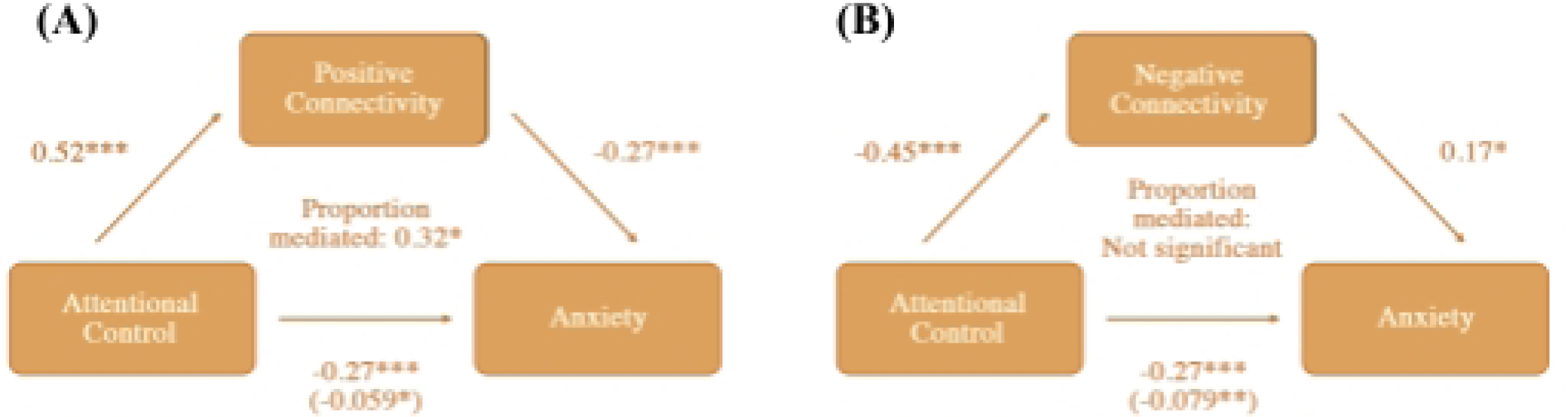
(A) Chord diagram depicting att.

### 3.2. Mediation Analysis

Attention control positively predicted anxiety and depression symptoms. Attention control also positively predicted ACPN and negatively predicted ACNN. ACPN negatively predicted both anxiety and depression symptoms, while ACNN positively predicted both anxiety and depression symptoms. These findings were all significant and are reflected in Table 1.

**Table 1.**
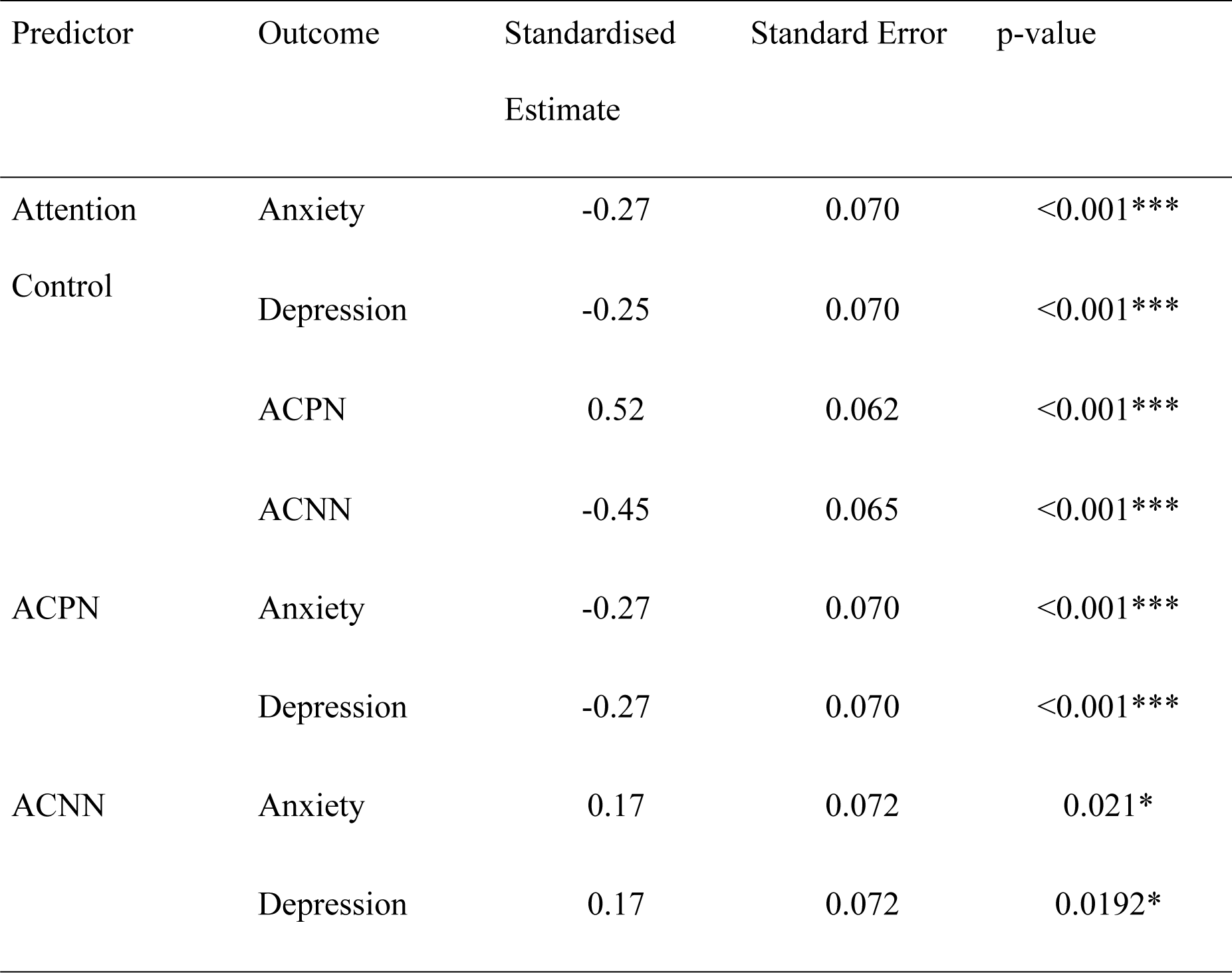
Summary of All the Regression Analyses Not Yet Accounting for the Mediator.

The effect of attention control on anxiety was found to be partially mediated via the ACPN but was not significantly mediated via the ACNN. Results of the mediation analysis for anxiety are shown in Table 2 and Figure 3. The effect of attention control on depression was fully mediated via the ACPN but was not significantly mediated via the ACNN. These results can be found in Table 3 and Figure 4.

**Figure 3.**
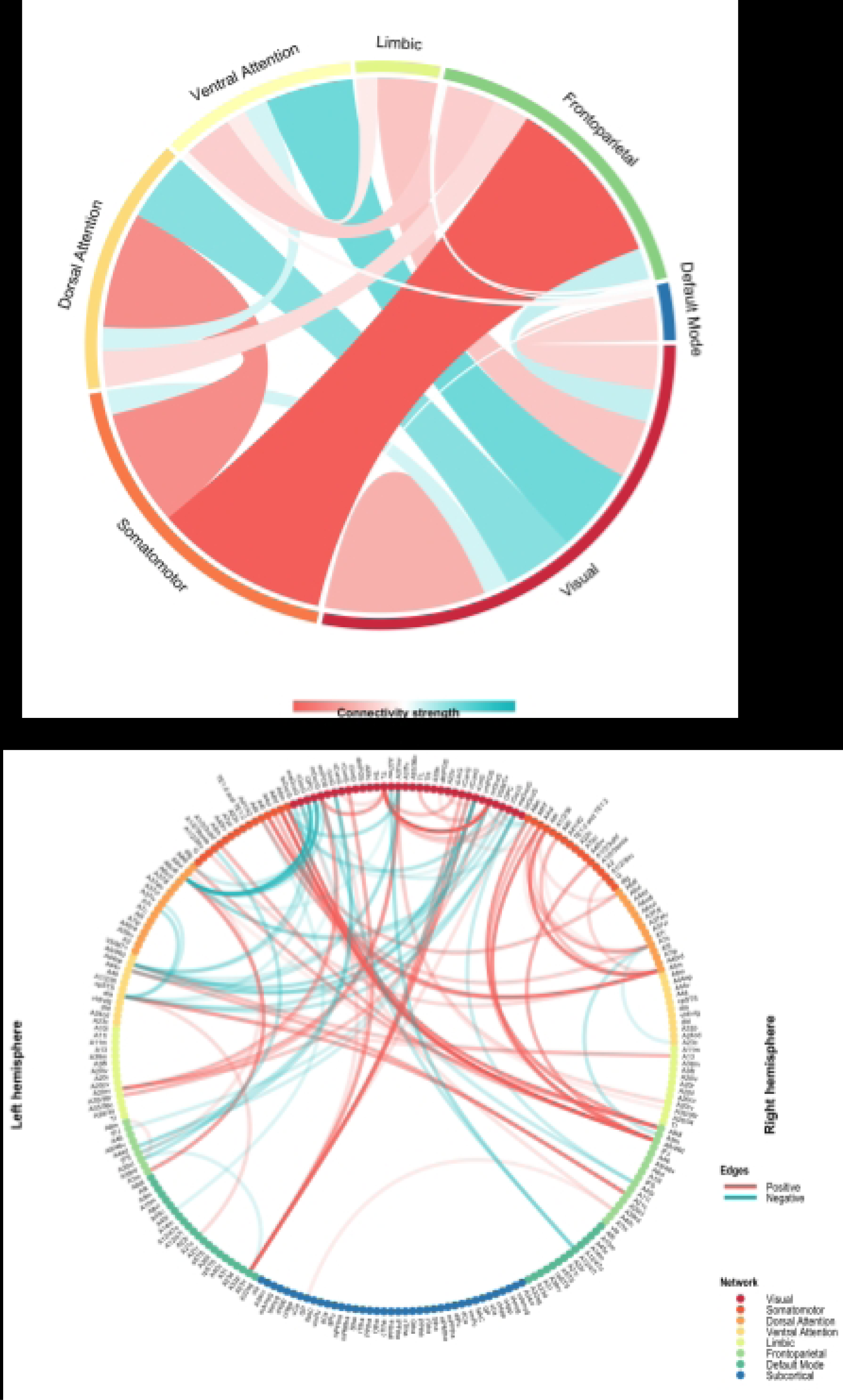
Visualisation of me.

**Figure 4.**
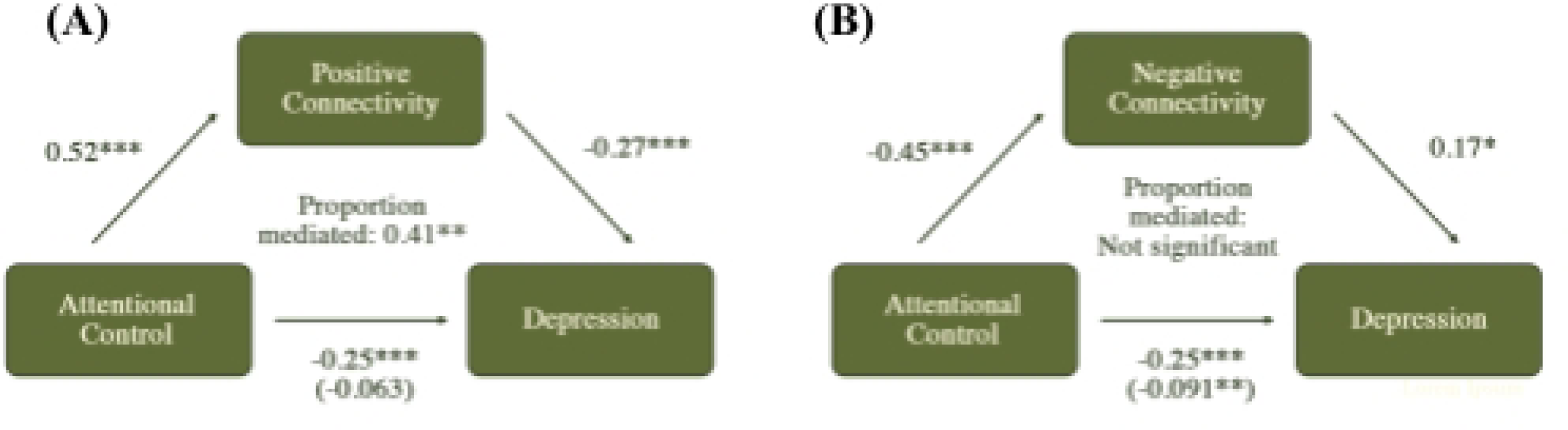
Visualisation of mediation moc.

**Table 2.**
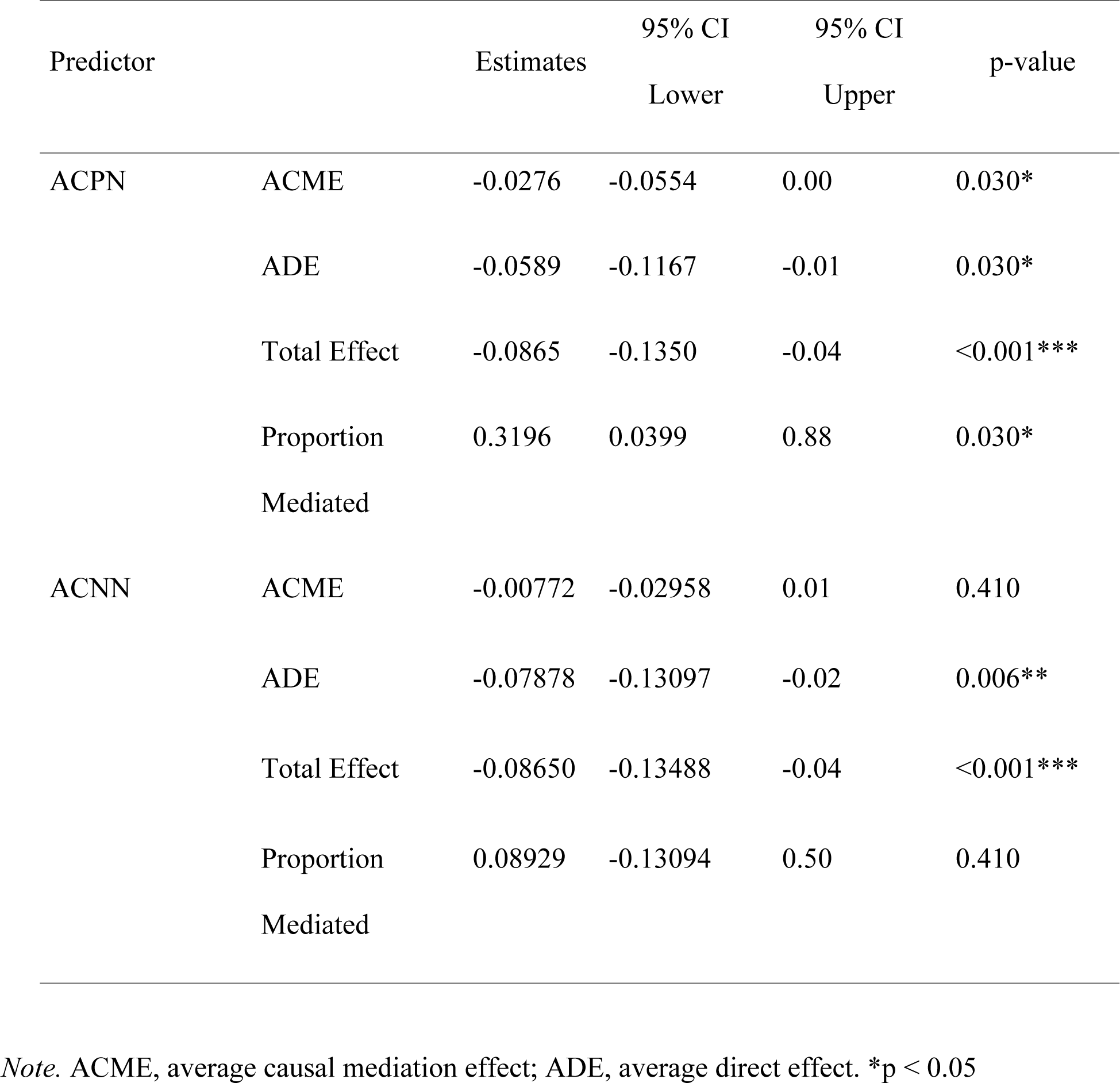
Estimates of the ACPN- and ACNN-mediated Model of Attention Control Against Anxiety.

**Table 3.** Estimates of the ACPN- and ACNN-mediated Model of Attention Control Against Depression.

| Predictor |  | Estimates | 95% CI<br>Lower | 95% CI<br>Upper | p-value |
| --- | --- | --- | --- | --- | --- |
| ACPN | ACME | -0.101 | -0.181 | -0.03 | 0.008** |
|  | ADE | -0.147 | -0.309 | 0.01 | 0.064 |
|  | Total Effect | -0.248 | -0.406 | -0.11 | <0.001*** |
|  | Proportion Mediated | 0.406 | 0.114 | 1.07 | 0.008** |
| ACNN | ACME | -0.0138 | -0.0412 | 0.01 | 0.34 |
|  | ADE | -0.0914 | -0.1591 | -0.02 | 0.01** |
|  | Total Effect | -0.1052 | -0.1682 | -0.05 | <0.001*** |
|  | Proportion Mediated | 0.1315 | -0.1509 | 0.56 | 0.34 |

*Note.* ACME, average causal mediation effect; ADE, average direct effect. \*p < 0.05

## 4. Discussion

This study identified rsFC positively and negatively associated with attention control. The pattern of edges in these networks reflected an increased somatomotor-frontoparietal and dorsal attention-somatomotor connectivity, and a decreased dorsal attention-visual and ventral attention-visual connectivity in attention control. The ACPN partially mediated the effect of attention control on anxiety and fully mediated the effect of attention control on depression. On the other hand, the ACNN did not significantly mediate the relationship between attention control and anxiety or depression.

What is surprising about the results is the increased connectivity between the frontoparietal network and dorsal attention network with the somatomotor network in attention control. The current literature on the role of the frontoparietal- and/or dorsal attention-somatosensory network on attention mostly comprises clinical groups with attention deficits, namely in ADHD populations. A mix of findings have been found, such as stronger connections in an ADHD population between the white matter rsFC of the default mode network and the somatomotor network with the other networks [62], increased visual-dorsal attention connectivity [63] and increased surfaced-based brain rsFC within the limbic, visual default mode, somatomotor, dorsal attention, frontoparietal and VANs [64]. Poor sustained attention in this population was also associated with stronger positive connectivity within the motor network bilaterally and between motor, parietal, prefrontal, and limbic networks in a task-based functional connectivity study [65]. Poor motor control has been observed in ADHD children, which may imply a lower somatomotor activity correlation with inattentiveness [66]. Other studies did look at the somatotopic (spatial) attention modulation of behaviour in non-clinical populations, but they were mainly limited to the neural processing of tactile stimuli [67–73, 104] and most do not investigate functional connectivity of the somatosensory region with other regions of the brain.

To the best of our knowledge, this paper is the first to look into the rsFC of attention control in all age groups and non-clinical populations. The implication of the somatomotor network in attention may have to do with the fact that the ACS measured a somatic component of attention, such as one’s ability to alternate between different modalities of external sensory input (e.g. “When I am reading or studying, I am easily distracted if there are people talking in the same room.”) or regulate internal sensory input (e.g. “When concentrating I ignore feelings of hunger or thirst.”). In comparison, the attention control measures in previous literature might not have paid as close attention to these somatic components. Some [74–76] used task-based attentional search assessments such as the ANT-I task [78] which had a heavier cognitive load but only required minor motor responses, while others [79] used questionnaires with less emphasis on the somatic components and more emphasis on the affective components of anxiety, such as the State Trait Anxiety Inventory which includes items such as “I worry too much over something that really doesn’t matter” and “I feel calm; I feel secure.”

Another interesting finding of this paper was that anxiety symptoms were not correlated with the frontoparietal- or dorsal attention-somatomotor rsFC strength. This paper’s findings hence dispute previous findings about the positive correlation of somatomotor connectivity with heightened anxiety [80–82] as increased rsFC of the frontoparietal- and dorsal attention-somatomotor network was found in this study to be associated with lowered anxiety levels. The findings of past studies mainly clustered around the decreased functioning within the frontoparietal network during the processing of neutral targets [75, 76, 79, 83] and increased activity in these portions in tasks that use emotionally laden stimuli [76, 84–86] in individuals high in trait anxiety or diagnosed with an anxiety disorder. Other studies focusing on the DAN tend to focus mainly on dorsal attention-amygdala activity [87–90]. A possible reason for this discrepancy is that some of the previous literature examined clinically anxious populations [85, 86] while this current study investigated rsFC in sub-clinical and non-clinical populations. However, for the rest of the studies that similarly focused on healthy individuals, the cause for this discrepancy could be a focus for future studies.

The last finding of this paper worth mentioning would be that while the HADS.A scale measured somatic-related anxiety symptoms (such as “I feel tense or ‘wound up.’”), the HADS.D scale does not measure any somatic symptoms related to depression. Yet, ACPN was found to be a partial mediator in the relationship between attention control and anxiety symptoms and a complete mediator in the relationship between attention control and depression symptoms. This may imply that the attention control networks play a more important role in this relationship. We propose that this may stem from the connection between rumination, known to impede attention control [91], and the potentially weakened connectivity within the dorsal attention and frontoparietal somatosensory networks. This reduced connectivity could hinder individuals from redirecting negative thought patterns and engaging in mindfulness-enhancing motor activities, thereby potentially exacerbating mood symptoms [92, 93]. Specifically, ruminative thoughts related to self-efficacy can intensify depression symptoms, while those concerning perceived social failures can heighten anxiety [92]. While rumination plays a key role in both anxiety and depression symptom perpetuation [94], it is a hallmark symptom of depression but only a potential symptom of anxiety [95]. Our current study aligns with the findings of [96], who identified hypoconnectivity between the resting-state functional connectivity (rsFC) frontoparietal and dorsal attention networks in Major Depressive Disorder (MDD). However, their study did not explicitly investigate the somatomotor network, likely due to its primary focus on MDD in general rather than exploring the link between attention control and depression specifically.

The findings of this study would be useful for neuroeducation, “a didactic or experiential-based intervention that aims to reduce client distress and improve client outcome by helping clients understand the neurological processes underlying mental functioning” [97]. In the context of this study, neuroeducation would involve informing clients of the neurological processes, strengthening connectivity in their frontoparietal- or dorsal attention-somatomotor network which may underlie their poor attention control. This knowledge empowers clients to develop self-compassion, modify deep-seated cognitive patterns, and normalise the fluctuations in their personal growth journey [97]. These insights have the potential to alleviate client anxiety, enhance results, and foster greater cooperation between therapists and their clients. [104]. Furthermore, the findings support therapies that incorporate somatic and attention-based elements, like mindfulness cognitive-based therapy, which has demonstrated effectiveness in reducing anxiety and depression symptoms such as rumination [93, 98–100] and enhancing emotion regulation abilities [101, 102].

The findings are subject to some limitations. Firstly, the present study is purely correlational. Causation between attention control, functional connectivity of the brain and anxiety- and depression-related symptoms cannot be inferred. Second, attention control was measured via self-reported questionnaire responses which may be subjective and unreliable compared to more objective measures of attention control, such as using a Stroop task. Replication of the present study with more objective measures or with additional measures for corroboration such as peer-reported measures could increase its validity.

## Data Availability

Data was not collected firsthand. The open-access anonymous database utilised in our study may be accessed via Babayan et al., 2019 and Mendes et al., 2019.

http://fcon_1000.projects.nitrc.org/indi/retro/MPI_LEMON.html.

## Author Contributions

Raye Fion Loh: Conceptualization, Formal analysis, Investigation, Project administration, Software, Visualization, Writing – original draft

Savannah Siew Kah Hui: Supervision, Visualization, Writing – review & editing

Junhong Yu: Data curation, Funding acquisition, Methodology, Resources, Software, Supervision, Writing – review & editing

